# Correlation between COVID-19 vaccination and inflammatory musculoskeletal disorders

**DOI:** 10.1101/2023.11.14.23298544

**Authors:** Young Hwan Park, Min Ho Kim, Myeong Geun Choi, Eun Mi Chun

**Author notes:** Corresponding author: Eun Mi Chun, MD, PhD, Division of Pulmonary and Critical Care Medicine, Department of Internal Medicine, Mokdong Hospital, Ewha Womans University College of Medicine, 1071 Anyangcheon-ro, Yangcheon-gu, Seoul 07985, Korea.

## Abstract

**Importance:** Earlier research on COVID-19 vaccines identified a range of adverse reactions related to proinflammatory actions that can lead to an excessive immune response and sustained inflammation. However, no study has been conducted on the association between inflammatory musculoskeletal disorders and COVID-19 vaccines.

**Objective:** To investigate the incidence rates of inflammatory musculoskeletal disorders following COVID-19 vaccination and to compare them with those of unvaccinated individuals.

**Design, Setting, and Participants:** This retrospective nationwide cohort study used data from the Korean National Health Insurance Service (NHIS) database, involving 2,218,715 individuals. Data were collected from January 1, 2021, to 12 weeks after the second dose of vaccine for vaccinated individuals and 12 weeks after September 30, 2021, for unvaccinated individuals.

**Exposures:** Status was categorized as unvaccinated and vaccinated with mRNA vaccine, viral vector vaccine, and mixing and matching.

**Main Outcomes and Measures:** The primary outcome was the occurrence of inflammatory musculoskeletal disorders that were selected as plantar fasciitis (ICD code, M72.2), rotator cuff syndrome (M75.1), adhesive capsulitis (M75.0), herniated intervertebral disc (HIVD) (M50.2/M51.2), spondylosis (M47.9), bursitis (M71.9), Achilles tendinitis (M76.6), and de-Quervain tenosynovitis (M65.4). Multivariate logistic regression analysis was used to determine the risk factors of musculoskeletal disorders after adjusting for potential confounders.

**Results:** Among the 2,218,715 individuals, 1,882,640 (84.9%) received two doses of the COVID-19 vaccine, and 336,075 (15.1%) did not. At 12 weeks after vaccination, the incidences of plantar fasciitis (0.14-0.17%), rotator cuff syndrome (0.29-0.42%), adhesive capsulitis (0.29-0.47%), HIVD (0.18-0.23%), spondylosis (0.14-0.23%), bursitis (0.02-0.03%), Achilles tendinitis (0.0-0.05%), and de-Quervain tenosynovitis (0.04-0.05%) were higher in all three vaccinated groups (mRNA, cDNA, and mixing and matching vaccines) when compared to the unvaccinated group. All COVID-19 vaccines were identified as significant risk factors for each inflammatory musculoskeletal disorder (odds ratio, 1.404-3.730), except for mixing and matching vaccines for de-Quervain tenosynovitis.

**Conclusions and Relevance:** This cohort study found that individuals who received any COVID-19 vaccine were more likely to be diagnosed with inflammatory musculoskeletal disorders than those who did not. This information will be useful in clarifying the adverse reactions to COVID-19 vaccines and informing people about their potential for inflammatory musculoskeletal disorders after vaccination.

## Introduction

The COVID-19 pandemic caused by the SARS-CoV-2 virus has had a profound impact on the world, with millions of infections and deaths reported globally. Therefore, extensive efforts were made to develop novel vaccines to combat the virus and curb its spread. The introduction of new COVID-19 vaccines has revolutionized vaccine science, offering unprecedented speed and efficacy during clinical trials.^1^ As a result, more than five billion people worldwide have been vaccinated so far, and declarations of public health emergencies have ended in most countries.^2^

Vaccines, inherently designed to stimulate the immune system, have the potential to elicit adverse reactions, which are often linked to immune-mediated responses involving vaccine excipients, active components, or immunodeficiency of the vaccinated individual.^3,4^ Traditional vaccines, with their longstanding accessibility to the public, have been associated with a predictable panel of adverse reactions, most of which are considered harmless.^3^ The COVID-19 vaccines are no exception to this; therefore, providing clear and transparent information about adverse reactions and demonstrating predictability are essential to increasing public trust and confidence.

Early research on COVID-19 vaccines identified a range of adverse reactions related to their proinflammatory effect, which could lead to an excessive immune response and sustained inflammation.^5^ However, other than presenting self-reported symptoms, no studies have been reported on the association of inflammation-related diseases with COVID-19 vaccines, especially in musculoskeletal disorders.^6–9^ Therefore, this study aimed to investigate the incidence of inflammatory musculoskeletal disorders after COVID-19 vaccination through a large-scale population survey. We used data from the Korean National Health Insurance Service (NHIS) database, which comprises a comprehensive dataset of 2,218,715 individuals.

## Methods

### Study Design and Population

This nationwide, population-based, retrospective cohort study used data from the Korean NHIS.^10^ On January 1, 2021, 50% of the residents of Seoul were randomly selected and included in the study population. The incidence of musculoskeletal disorders among the participants was then analyzed according to their vaccination status. Individuals who received two doses of the COVID-19 vaccine were defined as vaccinated, and their index date was the date of their second vaccination, prior to September 30, 2021. In contrast, the index date for unvaccinated individuals was September 30, 2021. Those who received only one dose of the vaccine and those who started vaccination after September 30, 2021, were excluded from the study. Diagnostic records for 365 d prior to the index date were reviewed, and individuals with any target musculoskeletal disorders as a primary or secondary diagnosis were excluded. If the target musculoskeletal disorder was the primary diagnosis on the day after the index date, it was defined as an event (Figure 1).

**Figure 1.**
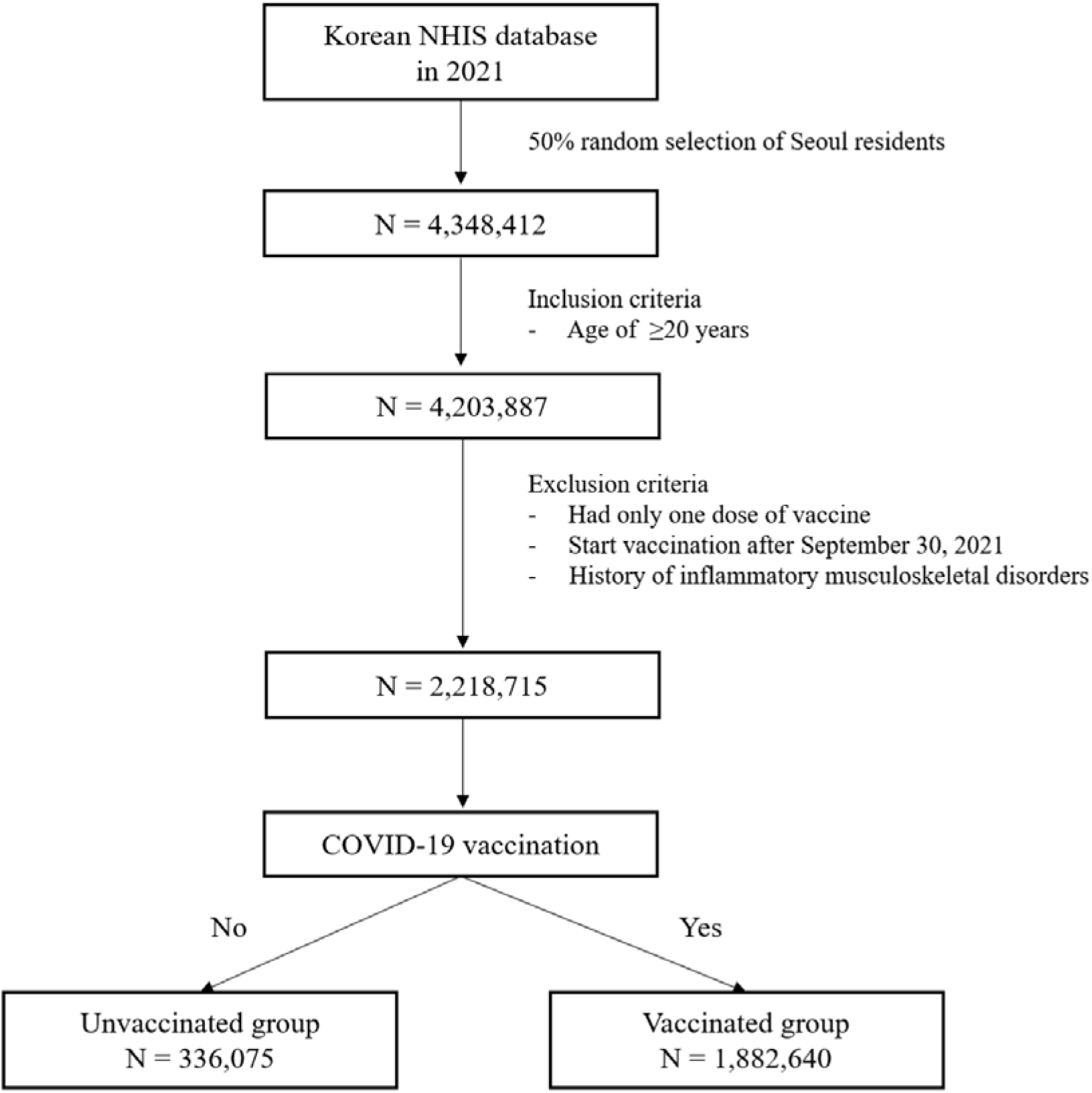
Flowchart of the study populations. Inflammatory musculoskeletal disorders include plantar fasciitis, rotator cuff syndrome, adhesive capsulitis, herniated intervertebral disc, spondylosis, bursitis, Achilles tendinitis, and de-Quervain tenosynovitis. NHIS, Korean National Health Insurance Service.

**Figure 2.**
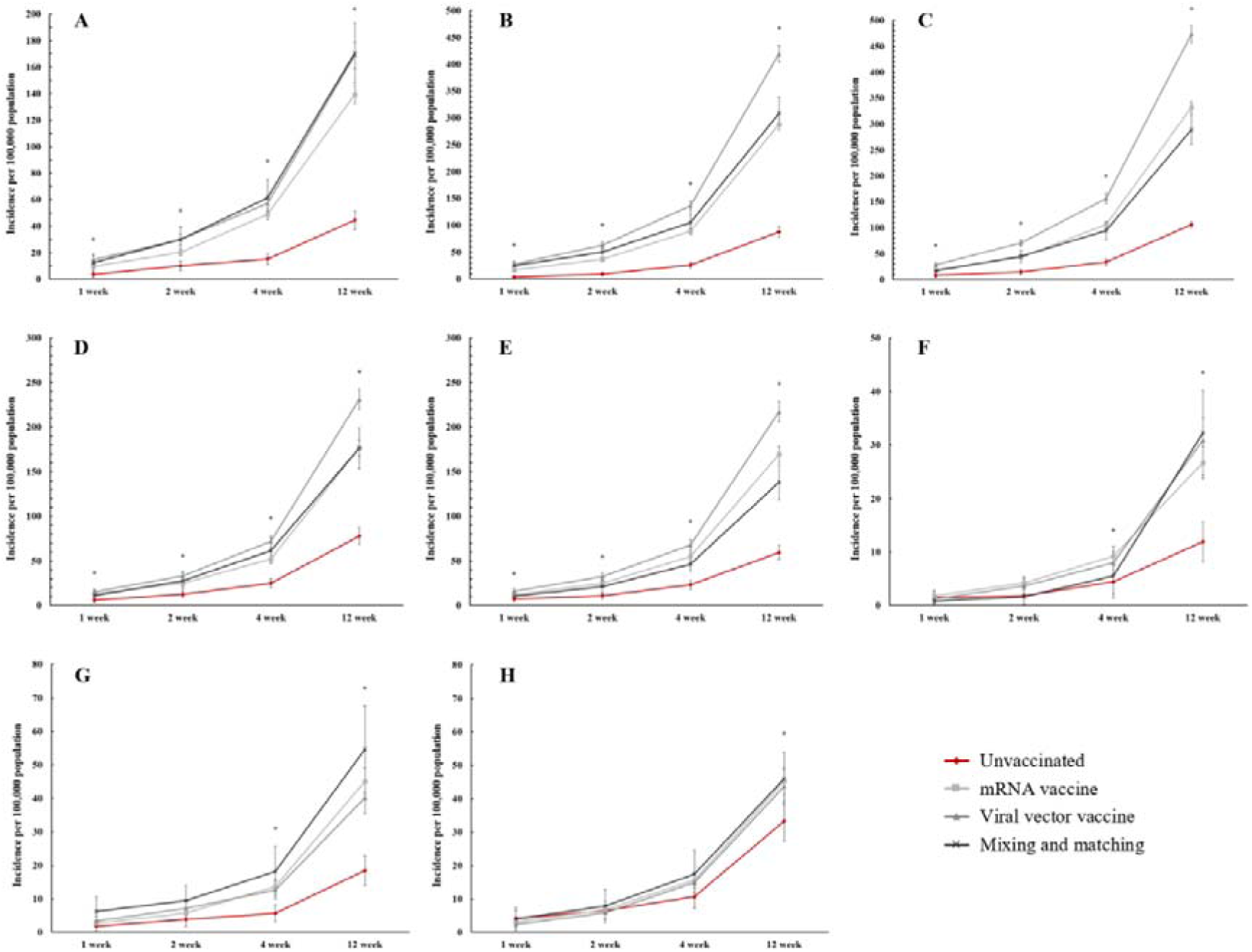
Incidence of inflammatory musculoskeletal disorders over time after COVID-19 vaccination. (A) plantar fasciitis; (B) rotator cuff syndrome; (C) adhesive capsulitis; (D) herniated intervertebral disc; (E) spondylosis; (F) bursitis; (G) Achilles tendinitis; and (H) de-Quervain tenosynovitis. Asterisks indicate that all three vaccinated groups are significantly higher than the unvaccinated group.

### Data Collection

We accessed the Korean NHIS data, which contains information on all medical claims, including diagnoses. The NHIS data were generated by the accumulation of claims by medical institutions under the Korean health insurance system.^11^ Data collection was approved by the NHIS and performed in accordance with the NHIS rules on data exploration and utilization.

Information on the demographic characteristics of the selected study population, such as age and sex, vaccination status and type, primary and secondary diagnoses from 2020 to 2021, were collected along with dates of hospital visits, underlying disease, and history of COVID-19. Age, sex, insurance level, Charlson comorbidity index (CCI), presence of diabetes, hypertension, hyperlipidemia, and chronic obstructive pulmonary disease (COPD), and history of COVID-19 were retrieved as covariates for the analysis. Regarding vaccination status, the participants were categorized as unvaccinated, messenger ribonucleic acid (mRNA)-vaccinated, viral vector-vaccinated, or mixed and matched. The mRNA vaccines were the Pfizer-BioNTech (BNT162b2) and Moderna (mRNA-1273) vaccines, while the viral vector vaccines were the Oxford-AstraZeneca (ChAdOx1 nCoV-19) and Johnson & Johnson (Ad26.COV2-S) vaccines. Mixing and matching vaccination was defined as the vaccination method that used heterologous vaccines in the first and second doses.^12^ The insurance level was classified as low, middle, and high based on the insurance premiums (medical benefit recipients and grades 1–6 are low, grades 7–13 are middle, and grades 14–20 are high). The CCI, which predicts the 10-year mortality of an individual with a range of comorbid conditions, was calculated based on a previous study by Sundararajan et al.^13,14^ The presence of diabetes, hypertension, hyperlipidemia, and COPD was defined as being registered as a primary or secondary diagnosis more than twice in the year prior to the index date. A history of COVID-19 infection was defined as the presence of the International Classification of Diseases 10th Revision (ICD-10) code U07.1, in either primary or secondary diagnoses before the index date.

The target musculoskeletal disorders that were selected to investigate their association with COVID-19 vaccination were musculoskeletal pathological conditions that could be induced, mediated, or aggravated by an inflammatory response. The selection of target musculoskeletal disorders was made through consensus among the authors based on previous studies, and the ICD-10 codes for search were as follows: plantar fasciitis (M72.2),^15^ rotator cuff syndrome (M75.1),^16,17^ adhesive capsulitis (M75.0),^18–20^ herniated intervertebral disc (HIVD) (M50.2/M51.2),^21–23^ spondylosis (M47.9),^24,25^ bursitis (M71.9),^26^ Achilles tendinitis (M76.6),^27,28^ and de-Quervain tenosynovitis (M65.4).^29,30^

### Ethical Approval

This study was approved by the local ethics committee and conducted in accordance with the ethical standards of the Declaration of Helsinki.^31^ The requirement for written informed consent was waived owing to the retrospective nature of the study.

### Statistical Analysis

The Student’s t-test was used to compare continuous variables, and the chi-square test or Fisher’s exact test was used to compare categorical variables. Incidence rates were calculated as rates per 100,000 individuals. With demographic characteristics of the study population, including the kind of vaccine received, multivariate logistic regression analysis was performed to determine the risk factors of inflammatory musculoskeletal disorders after adjusting for potential confounders. Associations between these factors and musculoskeletal disorders are summarized as odds ratios (ORs) and 95% confidence intervals (CIs). Statistical significance was set at p <0.05. The SAS Enterprise Guide (SAS Institute, Cary, NC, USA) was used for all statistical analyses and data curation.

## Results

Among the 2,218,715 individuals in the dataset, 1,882,640 (84.9%) received two doses of the COVID-19 vaccine, while 336,075 (15.1%) did not. The vaccinated group was older, predominantly female, had a higher insurance level, and had more comorbidities compared with the unvaccinated group (Table 1). Within the vaccinated group, the average time interval between the two vaccinations was 50.5 d. For each dose, 1,100,243 (58.4%) individuals received an mRNA vaccine, 656,184 (34.9%) received a viral vector vaccine, and 126,213 (6.7%) received a mixing and matching vaccine (Table 2).

**Table 1.** Demographic characteristics of the study population.

|  | Total | Vaccinated | Unvaccinated | p value |
| --- | --- | --- | --- | --- |
| Sex |  |  |  |  |
| Male | 1,017,422 (45.9) | 862,251 (45.8) | 155,171 (46.2) | <0.001 |
| Female | 1,201,293 (54.1) | 1,020,389 (54.2) | 180,904 (53.8) |  |
| Age |  |  |  |  |
| Mean ± SD, years | 54.1 ± 17.3 | 45.3 ± 17.2 | 55.7 ± 16.8 | <0.001 |
| 20–29 | 264,779 (11.9) | 201,896 (10.7) | 62,883 (18.7) | <0.001 |
| 30–39 | 241,993 (10.9) | 155,240 (8.3) | 86,752 (25.8) |  |
| 40–49 | 282,150 (12.7) | 211,572 (11.2) | 70,578 (21.0) |  |
| 50–59 | 508,893 (22.9) | 461,880 (24.5) | 47,016 (14.0) |  |
| 60–39 | 502,417 (22.7) | 468,525 (24.9) | 33,892 (10.1) |  |
| 70–79 | 276,783 (12.5) | 259,784 (13.8) | 16,999 (5.1) |  |
| $\geq 80$ | 141,700 (6.4) | 123,743 (6.6) | 17,957 (5.3) | |
| Insurance level |  |  |  |  |
| Low | 570,560 (25.7) | 472,383 (25.1) | 98,177 (29.2) | <0.001 |
| Middle | 611733 (27.6) | 506,536 (26.9) | 105,197 (31.3) |  |
| High | 1,036,422 (46.7) | 903,721 (48.0) | 132,701 (39.5) |  |
| Comorbidity |  |  |  |  |
| Diabetes mellitus | 351,517 (15.8) | 329,043 (17.5) | 22,474 (6.7) | <0.001 |
| Hyperlipidemia | 708,007 (31.9) | 666,137 (35.4) | 41,870 (12.5) | <0.001 |
| Hypertension | 624,705 (28.2) | 587,304 (31.2) | 37,398 (11.1) | <0.001 |
| COPD | 93,785 (4.2) | 85,634 (4.6) | 8,151 (2.4) | <0.001 |
| COVID-19 history | 18,411 (0.8) | 14,511 (0.8) | 3,900 (1.2) | <0.001 |
| CCI |  |  |  |  |
| 0 | 1,473,982 (66.4) | 1,194,333 (63.4) | 279,649 (83.2) | <0.001 |
| 1 | 372,684 (16.8) | 346,869 (18.4) | 25,815 (7.7) |  |
| ≥2 | 372,049 (16.8) | 341,438 (18.2) | 30,611 (9.1) |  |

Data are n (%), unless otherwise indicated.
SD, standard deviation; COPD, Chronic obstructive pulmonary disease; CCI, Charlson Comorbidity Index.

**Table 2.** Summary of the first and second doses of the COVID-19 vaccines received.

| First dose | Second dose | Number (%) |
| --- | --- | --- |
| Pfizer-BioNTech<sup>a</sup> | Pfizer-BioNTech | 1,072,100 (56.9) |
|  | Moderna | 21 (0.0) |
|  | Oxford-AstraZeneca | 4 (0.0) |
|  | Johnson & Johnson | 2 (0.0) |
| Oxford-AstraZeneca<sup>b</sup> | Oxford-AstraZeneca | 656,184 (34.8) |
|  | Pfizer-BioNTech | 126,196 (6.7) |
|  | Johnson & Johnson | 6 (0.0) |
|  | Moderna | 2 (0.0) |
| Moderna<sup>a</sup> | Moderna | 28,110 (1.5) |
|  | Pfizer-BioNTech | 5 (0.0) |
| Johnson & Johnson<sup>b</sup> | Oxford-AstraZeneca | 5 (0.0) |
|  | Pfizer-BioNTech | 5 (0.0) |

<sup>a</sup> mRNA vaccine
<sup>b</sup> Viral vector vaccine

The incidence of plantar fasciitis, rotator cuff syndrome, adhesive capsulitis, HIVD, and spondylosis was consistently higher in all three vaccination groups (mRNA, cDNA, and mixing and matching vaccines) than in the unvaccinated group. This disparity was evident at 2, 4, and 12 weeks after vaccination, with differences in incidence increasing over time (Figures 1-A, 1-B, 1-C, 1-D, and 1-E). For bursitis and Achilles tendinitis, no significant differences in incidence were observed between the three vaccinated groups and the unvaccinated group after 1 and 2 weeks of vaccination. However, at 4 and 12 weeks after vaccination, the incidences of bursitis and Achilles tendinitis were higher in the three vaccinated groups than in the unvaccinated group (Figures 1-F and 1-G). Up to 4 weeks after vaccination, no significant differences in the incidence of de Quervain tenosynovitis were observed; however, at 12 weeks after vaccination, the vaccinated group exhibited a significantly higher incidence than that of the unvaccinated group (Figure 1-H).

Multivariate logistic regression analysis showed that 6 weeks after vaccination, the mRNA vaccine, viral vector, and mixing and matching vaccines were significant factors for each musculoskeletal disorder, except for bursitis and de-Quervain tenosynovitis. Over a 12-week period, all vaccines showed statistical significance in relation to each musculoskeletal disorder, except for the mixing and matching vaccines for de-Quervain tenosynovitis (Table 3). In addition, female individuals and older individuals were vulnerable to musculoskeletal disorders other than bursitis and Achilles tendinitis after COVID-19 vaccination (Supplement 1).

**Table 3.**
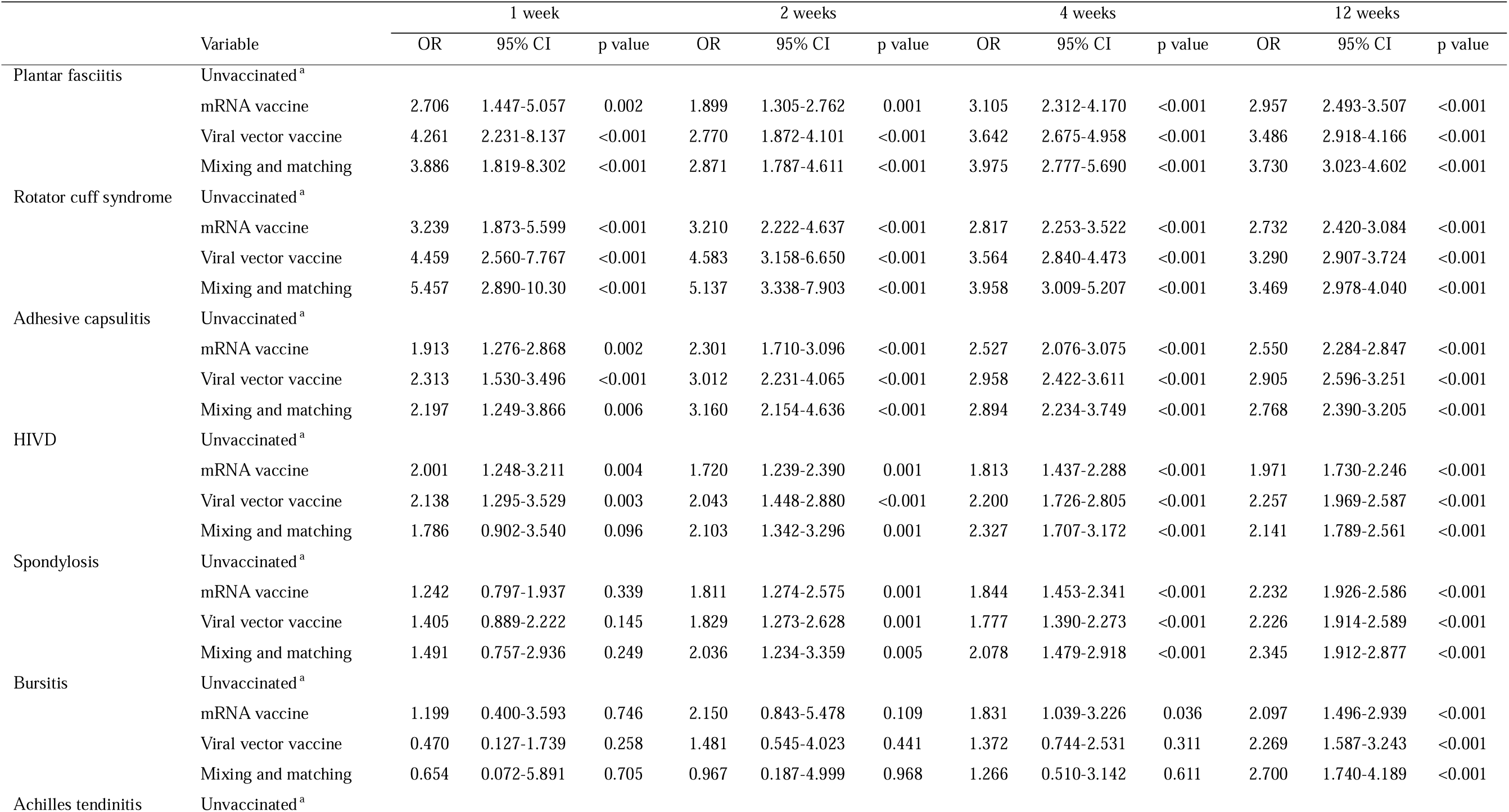
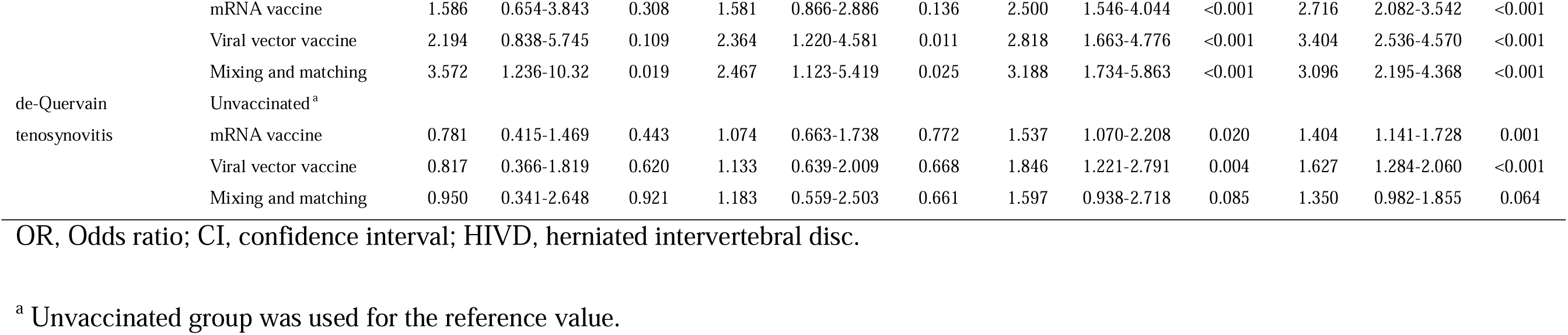
Results of multivariate logistic regression model to assess the risk of musculoskeletal disorders after COVID-19 vaccination.

## Discussion

In this nationwide population-based study, the incidence rates of inflammatory musculoskeletal disorders in terms of adverse reactions after COVID-19 vaccination in Korea were investigated. We found that initially after vaccination, the incidences of plantar fasciitis, rotator cuff syndrome, adhesive capsulitis, HIVD, and spondylosis, which are inflammation-related disorders involving tendons and connective tissues, were higher in the vaccinated group than in the unvaccinated group, regardless of the type of vaccine received. At 12 weeks after vaccination, all inflammatory musculoskeletal disorders investigated, including bursitis, Achilles tendinitis, and de Quervain tenosynovitis, showed a higher incidence in the vaccinated group than in the unvaccinated group. These findings provide detailed information on the adverse reactions after COVID-19 vaccination, especially in terms of inflammation-related musculoskeletal disorders. We believe that this information will be beneficial for establishing the predictability of adverse reactions and enhancing public understanding of COVID-19 vaccination.

Inflammatory musculoskeletal manifestations after COVID-19 vaccination have been repeatedly reported in case reports and small cohort studies.^32^ In one of the largest case studies conducted, Ursini et al. concluded that inflammatory musculoskeletal disorders may occasionally develop in close temporal association with COVID-19 vaccination.^33^ Along with these previous studies, our study highlights the development of inflammatory musculoskeletal disorders after COVID-19 vaccination. However, an association between COVID-19 vaccines and inflammatory musculoskeletal disorders is yet to be established. According to the hypothetical mechanisms of autoimmune phenomena after COVID-19 vaccination, either adjuvants or the vaccine itself may induce an overactive immune reaction, autoimmune consequences, or even inflammation in susceptible individuals.^34^ As a form of molecular mimicry, a hypothesis states that overwhelming systemic or local inflammation is activated by the cross-reaction of the immune response between antigens in vaccines and molecular structures *in vivo*.^35^ The series of inflammatory musculoskeletal disorders that showed a strong correlation with COVID-19 vaccinations in this study are expected to be included in the list of adverse reactions to COVID-19 vaccines. Therefore, future studies are needed to identify the mechanisms that link COVID-19 vaccines to inflammatory musculoskeletal disorders and prevent or mitigate these adverse reactions.

Regarding the inflammatory musculoskeletal disorders investigated in this study, the incidence rates of shoulder-related disorders, such as rotator cuff syndrome and adhesive capsulitis, were notably higher than those of other disorders—nearly doubling the rate of other disorders. We believe that shoulder injury related to vaccine administration (SIRVA),^36^ which has already been reported for existing vaccines, may have contributed to the high incidence rates observed. SIRVA is thought to occur by provoking an inflammatory reaction when vaccines are injected through the deltoid into underlying non-muscular tissues.^37^ Subsequent prolonged inflammatory responses after SIRVA could progress to bursitis, tendinitis, and capsulitis around the shoulder joint.^38^ The findings of this study indicate that the COVID-19 vaccine is not exempt from SIRVA. Therefore, as with other vaccination procedures, we emphasize increased attention to the injection site during the administration of the COVID-19 vaccine.

Previous studies on adverse reactions to the COVID-19 vaccine showed that most systemic adverse reactions were more frequent in female individuals than in male individuals and in younger age groups than in older age groups.^6,7^ In our study, the incidence rates of inflammatory musculoskeletal disorders, regarded as adverse effects, were higher in females, which is consistent with the reports of previous studies. However, in terms of age, older individuals had a higher incidence of inflammatory musculoskeletal disorders compared to younger individuals. Two possible explanations exist for the inconsistency between our age-specific findings and those of previous reports. First, older individuals exhibit a reduced capacity to mount an effective response to vaccines as well as a lower frequency of neutralizing antibodies compared to younger populations.^39,40^ Moreover, older patients may be more prone to progressing from mild adverse reactions to inflammation-related musculoskeletal disorders compared to their younger counterparts. Similarly, although the overall incidence of adverse reactions after COVID-19 vaccination was higher in the younger age group, the incidence of severe adverse reactions was reported to be higher in the older age group.^41^ Second, as with other adverse reactions, inflammatory musculoskeletal disorders occurred more frequently in the younger than in the older age group; however, in the dataset, diagnosis and insurance claims appeared to occur more frequently in older individuals because younger patients visited medical institutions less often. Irrespective of which of these two possibilities is true, we maintain that the need for attention after vaccination in older individuals remains unchanged.

### Strengths and Limitations

The strength of our study is underscored by its substantial sample size, which comprises data from over 2 million individuals randomly selected from the Korean NHIS. This comprehensive database, which encompasses medical services for 97% of the population, enhances the reliability and representativeness of our findings.^10^ Such large population-based databases, which are available only in Taiwan, Sweden, and Korea, are excellent resources for answering questions that are difficult to address using single-institution or small-scale studies.^42^

Our study also has several limitations. First, because the target musculoskeletal disorders were identified by ICD-10 codes in the claims database, coding, mismatching, or misclassification errors could have occurred. Discrepancies might have occurred between the actual disease and the diagnosis claimed by the healthcare provider, and over- or underdiagnosis may occur. Second, our study could not confirm the pathophysiological mechanism of the change in the incidence of musculoskeletal disorders after COVID-19 vaccination because it relied only on diagnoses claimed by healthcare providers. As COVID-19 vaccines are frequently accompanied by arthralgia and myalgia,^6,7^ whether these adverse reactions were misdiagnosed as inflammation-mediated musculoskeletal disorders or whether the pain was caused by an actual inflammatory disorder is unknown. Further studies involving laboratory data and inflammatory biomarkers are required to address these limitations.

## Conclusions

Individuals who received COVID-19 vaccines, either mRNA, viral vector, or mixing and matching, were found to be more likely to be diagnosed with inflammatory musculoskeletal disorders compared to those who did not. Our results provide detailed information on the adverse reactions after COVID-19 vaccination, with a particular focus on inflammatory musculoskeletal disorders. This information will be useful in clarifying adverse reactions to COVID-19 vaccines and educating people about the potential risk of inflammatory musculoskeletal disorders based on their vaccination status.

## Data Availability

All data produced in the present study are available upon reasonable request to the authors.

## Acknowledgment

None

## Data availability

See Appendix 1.

## Author Contributions

Dr Chun had full access to all of the data in the study and takes responsibility for the integrity of the data and the accuracy of the data analysis. Drs Park and Kim contributed equally as co–first authors.

Concept and design: All authors.

Acquisition, analysis, or interpretation of data: Park, Kim.

Drafting of the manuscript: All authors.

Critical revision of the manuscript for important intellectual content: Park, Chun.

Statistical analysis: Kim.

Obtained funding: None.

Administrative, technical, or material support: Park, Chun.

Supervision: Chun.

